# Antimicrobial stewardship among Nigerian children: A pilot study of the knowledge, attitude, and practices of prescribers at two tertiary healthcare facilities in Bayelsa State

**DOI:** 10.1101/2021.11.30.21267070

**Authors:** Ebiowei S.F Orubu, Faith O. Robert, Leonard Emuren, Boboye Ifie-Ombeh

## Abstract

Antimicrobial stewardship (AMS), the evidence-based use of antimicrobials, is an effective strategy in controlling antimicrobial resistance (AMR) in humans by reducing the irrational use of antimicrobials. Stewardship in children is less studied. This study assessed the knowledge, attitude, and practice of physicians prescribing antibiotics to children in Bayelsa State, Nigeria to identify gaps in AMS and possible solutions. Following ethical approval, a semi-structured questionnaire was distributed among 40 paediatricians and gynaecologists at the two public tertiary healthcare facilities in Bayelsa State – the Niger Delta University Teaching Hospital and the Federal Medical Centre – for self-completion. Responses were expressed as percentages and analyzed using Bloom’s cutoffs. The Capability, Opportunity, Motivation, and Behaviour (COM-B) model was employed to identify gaps for intervention in prescribing behavior with gaps in each component identified by aggregate scores <80%. Perceived approaches to improve prescribing among 14 selected options were assessed using 5-point Likert scales and options with scores >90% rated the most acceptable. Questionnaires were administered from August to September 2021. The response rate was 68% (27/40). Participants were paediatricians (81%, 22/27) and gynaecologists (19%, 5/27). Antimicrobial Susceptibility Testing (AST) was not performed before antibiotic selection nine times out of 10 (89%, 24/27). In a third (37%, 10/27) of cases, 2-3 antibiotics were prescribed. The top three antibiotics, in rank order, were: cefuroxime or amoxicillin 41% (11/27); ciprofloxacin or amoxicillin 30% (8/27), and azithromycin (33%, 9/27). Aggregate COM-B scores were: capability, 74%; opportunity, 78%; and motivation, 87%. The most acceptable (100%, 27/27) options to improving antibiotic prescribing were: availability of resistance data, availability of guidelines, readily accessible microbiological data, and easy access to infectious disease physicians. There are gaps in knowledge of AMR and opportunity for rational prescribing. There is need for antimicrobial resistance data to promote pediatric AMS at the surveyed healthcare facilities.

## Introduction

Antimicrobial resistance (AMR), an inherent and developed ability of microorganisms to withstand treatment with antimicrobial agents (1), is a global public health emergency (2,3). The factors contributing to the emergence and spread of AMR in humans are related to the misuse, underuse, or abuse of antimicrobial agents in humans, animals, agriculture, and the environment (1,4). Antimicrobial stewardship (AMS), **the evidence-based use of antimicrobials**, is an effective strategy in controlling AMR in humans by reducing the excessive use of antimicrobials (5). Overall, 50% of global antibiotic use is inappropriate, calling for urgent antimicrobial stewardship to reduce indiscriminate and excessive use of antibiotics (6).

Antimicrobial use in children tends to be higher than in adults, especially in developing countries (7). In most Low- and Middle-Income Countries (LMICs), the high burden of infectious diseases, unrestricted access to antimicrobials, and poor awareness promote the indiscriminate use of antimicrobials and drive AMR (8–12). Emerging research show that the use of substandard and falsified (SF), or poor-quality, antimicrobials can also contribute to AMR (13,14).

Nigeria, together with four other LMICs, leads the world in deaths from infectious diseases in young children (<5 years old), mostly from lower respiratory tract infections (pneumonia), diarrhea, malaria, and perinatal and neonatal infections. There is little literature on antimicrobial use and antimicrobial stewardship programs in the paediatric population in Nigeria. There are gaps in antimicrobial stewardship activities, generally for Nigeria and specifically for children. A review of the literature on AMS in Nigeria failed to show any studies on AMS in children (15–21).

This study aims to understand providers’ knowledge, attitude, and practice on antimicrobial use, resistance, and stewardship with the ultimate goal of designing an AMS program for children at the tertiary healthcare level in Nigeria. The objectives were to: (i) assess current antimicrobial prescribing behaviour; and (ii) identify acceptable options for improving rational antimicrobial prescribing for children in this setting.

## Method

Study design: Cross-sectional questionnaire survey.

Ethics: Ethical approval for the study was obtained from NDUTH (NDUTH/REC/0040/2021).

Study location: The study location was Bayelsa State, one of the 37 federating units of 36 states and the Federal Capital Territory in Nigeria (Figure 1). It was conveniently selected.

**Figure 1.**
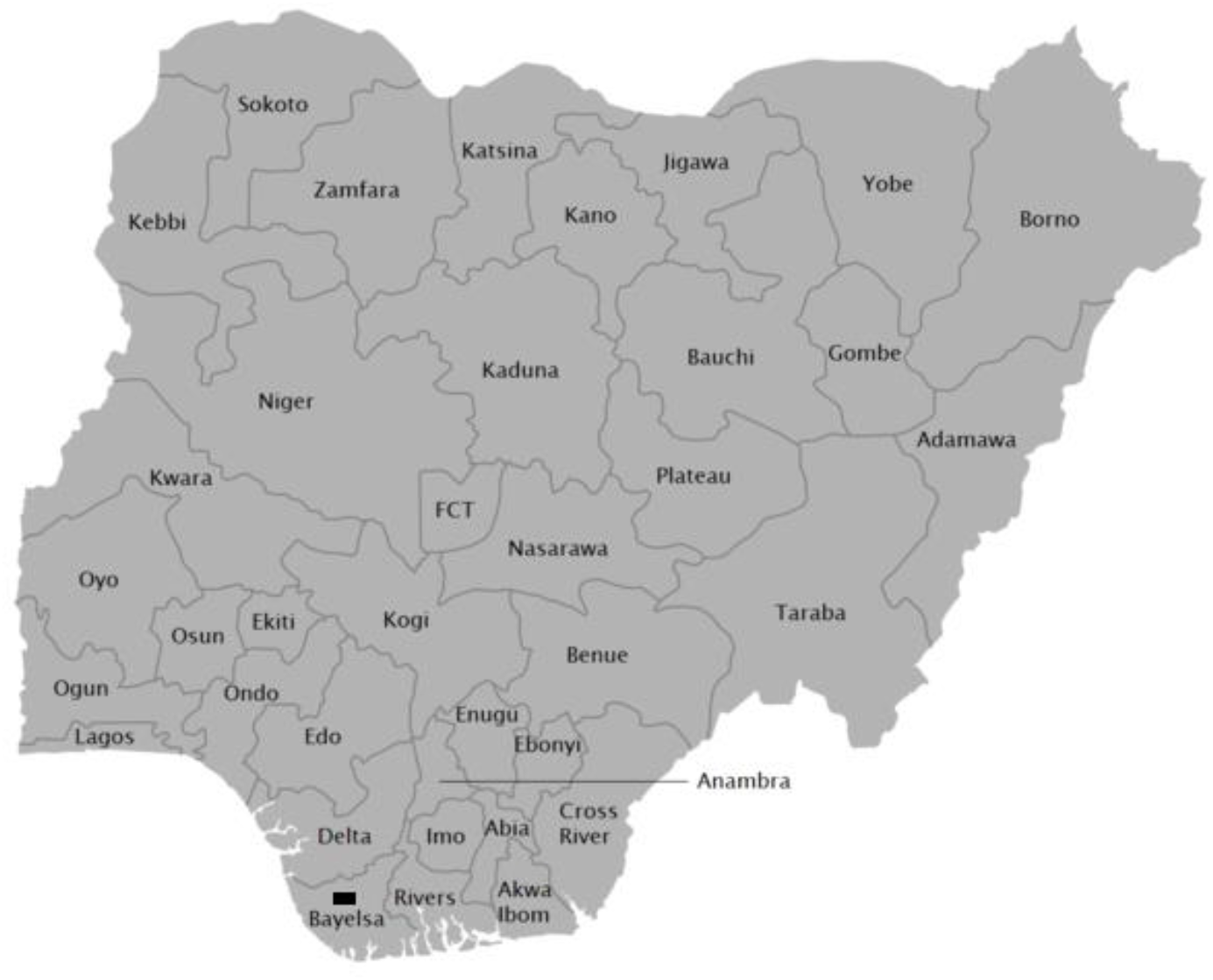
Map of Nigeria with study location indicated by the rectangle. [Source:https://en.wikipedia.org/wiki/File:Map_of_Nigerian_States_with_names.png#/media/File:Map_of_Nigerian_States_with_names.png

Study setting: The study settings were the two public tertiary-level healthcare facilities in Bayelsa State: the Niger Delta University Teaching Hospital (NDUTH), Okolobiri, the primary study location, and the Federal Medical Centre (FMC), Yenagoa. The NDUTH is the teaching hospital of the Niger Delta University. It is a 200-bed hospital providing comprehensive general and specialist healthcare for adults and children (22). Its paediatric department has 25 beds and 30 paediatricians. The FMC is a 425-bed hospital with a paediatrician staff strength of 50.

Sample size: A sample size of 20 pediatricians was targeted, generally considered appropriate for a pilot study (23,24). To provide information on prescribing practices around pregnancy and childbirth, the survey was extended to prescribers with obstetrics and gynaecology (O&G) specialization, increasing the sample size to 40.

Participants: Participants were purposively selected as pediatricians and obstetrics and gynecology specialists.

Study instrument: The survey instrument was a semi-structured questionnaire adapted from previously used and validated questionnaire surveys assessing knowledge, attitude, and practice of prescribers and other health workers in Europe (25,26) and Yemen (27). The questionnaire was arranged in 3 “sections” and contained 40 items, not counting an initial consent question and asking any additional question at the end (Supplement Table 1). The questions were adaptive, such that participants did not have to answer all but answered related questions based on their selection of a previous option. Selected questions were repeated to check for individual response reliability. Section I collected demographic data on the prescriber’s specialty and years of experience (2 questions). Section II assessed knowledge, attitude and practice, or prescribers’ behaviour. It contained 37 questions on current antibiotic prescription patterns and decision-making, knowledge of, and attitude or perceptions, about both AMR and existing antimicrobial stewardship structures. Knowledge was both real (assessed by 1 question containing 7 “key” statements with true or false answers) and assumed, or perceived, (assessed by 3 questions with Yes/No answers or 5-point Likert scales). Section III consisted of one question that further evaluated perceptions among 14 options for improving antibiotic prescribing, or AMS. The questionnaire was made available in both paper and electronic forms. The online questionnaire was prefaced by an introductory section that presented the title, aims, and objectives of the study in an information section and consent, while the paper form included separate information and consent form sections. To ensure completeness in filling the online questionnaire, responses to the main questionnaires were made necessary for submission.

Questionnaire administration: Questionnaires were distributed for self-administration in August 2021. One researcher (BIO) distributed the paper questionnaire during a departmental meeting at NDUTH and shared the online link via social media (WhatsApp) informally through her professional network. Only participants who consented completed the study. All responses were de-identified.

### Analysis

Analysis was performed using descriptive statistics expressed in proportions or percentages. Questionnaires completed by hand were converted to digital forms for data analysis. Dummy variables were created for questions not completely filled in the paper forms. These were usually “false”, “unsure”, or assigned a “neutral” score. This applied to 5 questions in 2 otherwise completely filled paper questionnaires.

To characterize providers’ prescribing behaviour and identify gaps for interventions, the Capability, Opportunity, and Motivation for behavioural change, COM-B, model (26) was utilized (Supplement Table 2). Capability was an umbrella term encompassing individual knowledge, awareness, and perceptions. Opportunity was defined both as factors that lie outside the individual, including awareness or perception of existing facility-level AMS structures, as well as attitudes including confidence in prescribing. Motivation was all the brain processes that energize and direct behaviour and included a self-belief of a key role in the containment of AMR and association between prescribing and the emergence of AMR. All responses on a 5-point Likert scale were re-coded such that “strongly agree” and “agree” were assessed as “agree”, and “strongly disagree”, “neither agree nor disagree”, and “disagree” re-coded as “disagree”. Incomplete responses on paper were assigned the ‘neutral’ “neither agree nor disagree”. Bloom’s cutoffs were then applied to aggregate scores for the relevant responses to identify gaps to target for each behaviour component. With responses assessing participants’ knowledge and awareness of AMR, as well as opportunity for AMS, the cutoffs and interpretations were set at: >80% as “**good**”, 60-79% as “**moderate**”, and <60% as “**poor**” (28). For attitude, these cutoffs were, respectively, analyzed as “**positive**”, “**neutral**”, and “**negative**” (28). Motivation was assessed as “high” >80%) or “low” (≤80%). **Gaps were scores <80% in any one domain**.

To assess acceptable options for improving AMS, responses were re-coded and rated based on percentages. Responses on the 5-point Likert scale were aggregated with very helpful and helpful classified as “helpful” and neutral, unhelpful and very unhelpful re-coded as “unhelpful”. The options recategorized as helpful were then rated as “most acceptable” if they had aggregated scores >90%, “acceptable” with scores of >80-90%, and “maybe **acceptable**” if rated as helpful by 70-80% of respondents.

## Results

Responses are detailed in Supplement Table 3.

### I. Demographics

Response rate: The questionnaire was distributed to 40 prescribers, out of which 28 responded and completed the questionnaire, giving a participation, and response, rate of 70%. The response from one participant who did not belong to the two target specialties was excluded giving an actual, or adjusted, response rate of 68% (27/40).

Participants: Participants were paediatricians 81% (22/27) and gynaecologists 19% (5/27). The majority (85%, 23/27) had more than 5 years experience (37% with 6-10 years; 26% with 11-15 years; and 22% with >15 years’ experience) with only 15% (4/27) having less than 5 years experience. The majority (78%, 21/27) saw less than 25 patients a day, most of whom were children under 12 years old (Supplement Table 3a).

### II. Antibiotic prescribing practice/behaviour

#### Decision making

All participants (100%, 27/27) had prescribed an antibiotic within the three months preceding the survey. The primary considerations for prescribing antibiotics were the clinical severity of the case and symptoms the patient presented with (82%, 22/27, respectively). None rated patient demand as a reason for prescribing. A slight majority of prescribers (56%, 15/27) felt pressure to prescribe antibiotics. The main reasons for this pressure were the condition of the patient (100%, 15/15) and the fear of complications (93%, 14/15). Forty percent (6/15), agreed or strongly agreed that patient (caregiver/parent) demand was a reason for this pressure (Supplement Table 3b).

#### Pattern

The top three antibiotics usually prescribed were: amoxicillin (including amoxicillin-clavulanate) or cefuroxime which was the first choice of 41% (11/27), ciprofloxacin or amoxicillin (including amoxicillin-clavulanate) as the second choice by 30% (8/27), and azithromycin as the third choice (33%, 9/27). In slightly more than a third (37%, 10/27) of cases 2-3 antibiotics were prescribed.

Antibiotic classes most frequently used for Respiratory Tract Infections were penicillins 82% (22/27) and macrolides 78% (21/27). Fluoroquinolones were more frequently used for both Urinary Tract Infections and Gastrointestinal tract (GIT) diseases (63%, 17/27, and 66%, 18/27, respectively). Cephalosporins (56%, 15/27) and penicillins (44%, 12/27) were preferred for skin and soft tissue infections (Supplement Table 3c).

Seventy-four percent (20/27) prescribed generic medications, with the remaining 26% (7/27) prescribing by brand names. Quality was the main reason for prescribing by brand (100%, 7/7).

#### Guidelines

Standard Treatment Guidelines, or a guide specific to prescribing, was used by slightly more than half (55%, 15/27) of participants to guide antibiotic prescribing in their facility.

#### AST

Antimicrobial Susceptibility Testing (AST), the use of microbial culture and sensitivity tests, were not performed before antibiotic selection nine times out of 10 (89%, 24/27). This was even the case when there was the possibility (67%, 18/27) of performing AST on-site (100%, 18/18). The most common barriers to AST were time pressure (79%, 19/24) and/or availability of reagents or materials (50%, 12/24).

#### Treatment failure rates

Treatment failure in patients within a month was perceived by the majority (96%, 26/27) of respondents as ranging from <10% (by 52%, 14/27) to between 26-50% (by 11%, 3/27).

### III. Capability, Opportunity and Motivation for Behaviour Change

AMR capacity: The knowledge and ability to inform others about AMR and use this to guide appropriate prescribing ranged from 70% (moderate) to 89% (good) (Table 1). Overall, AMR capacity was good (81%).

**Table 1:** Knowledge of AMR and the ability to inform others and to use this to guide prescribing ranked by the re-coded “agree”

|  |  | **Strongly Disagree** | **Disagree** | **Neither disagree nor agree** | **Agree** | **Strongly agree** | **"Agree"** |
| --- | --- | --- | --- | --- | --- | --- | --- |
| **a** | I know what antibiotic resistance is | 3 |  |  | 6 | 18 | 89% |
| **b** | I know what information to give to individuals about prudent use of antibiotics and antibiotic resistance | 3 |  | 1 | 5 | 18 | 85% |
| **c** | I know sufficient knowledge about how to use antibiotics appropriately for my current practice | 2 | 3 | 3 | 7 | 12 | 70% |
| **Aggregate score** |  |  |  |  |  |  | **81%** |

Knowledge: There were individual variations in answering the 7 “key” knowledge statements. Only two of these were correctly answered by all 27 (100%). To varying degrees, the answers to the other 5 on knowledge of decreased efficacy of antibiotics with unnecessary use, potential side effects, increased potential of AMR, transmissibility between people, and carriage of AMR in healthy individuals had correct answers ranging from 70-96%. The two most incorrectly answered questions were: “Healthy people can carry antibiotic-resistant bacteria” (70% [19/27], yes) and “Every person treated with antibiotics is at an increased risk of antibiotic-resistant infections” (74% [20/27], yes) (Table 2). Aggregate real knowledge was good.

**Table 2:** Knowledge about the dynamics of AMR emergence and spread rated by the overall proportion of correct answers

|  |  | **TRUE** | **FALSE** | **Correct answer** | **Correct (%)** |
| --- | --- | --- | --- | --- | --- |
| **a** | Antibiotics are effective against viruses |  | 27 | FALSE | 100% |
| **b** | Antibiotics are effective against cold and flu |  | 27 | FALSE | 100% |
| **c** | Taking antibiotics has associated side effects such as diarrhea, colitis, or allergy | 26 | 1 | TRUE | 96% |
| **d** | Unnecessary use of antibiotics makes them become ineffective | 25 | 2 | TRUE | 93% |
| **e** | Antibiotic resistant bacteria can spread from person to person | 22 | 5 | TRUE | 81% |
| **f** | Every person treated with antibiotics is at an increased risk of antibiotic resistant infection | 20 | 7 | TRUE | 74% |
| **g** | Healthy people can carry antibiotic resistant bacteria | 19 | 8 | TRUE | 70% |
| **Aggregate score** |  |  |  |  | **88%** |

One Health: Knowledge of One Health contributors to AMR varied widely. Only 37%, 11/27) correctly agreed that environmental factors such as wastewater contributes to AMR. In contrast, 70%, 19/27) correctly agreed that the excessive use of antibiotics in livestock and food production contributes to AMR (Table 3). The aggregate One Health knowledge score was poor.

**Table 3.** Knowledge about One Health components of AMR

|  |  | **Strongly**<br>**Disagree** | **Disagree** | **Neither**<br>**disagree**<br>**nor**<br>**agree** | **Agree** | **Strongly**<br>**agree** | **"Correct"** |
| --- | --- | --- | --- | --- | --- | --- | --- |
| **a** | Environmental factors such as waste water in the environment | 2 | 3 | 12 | 7 | 3 | 37% |
| **b** | Excessive use of antibiotics in livestock and food production | 1 | 1 | 7 | 14 | 4 | 67% |
| **Aggregate score** |  |  |  |  |  |  | **52%** |

Awareness of WHO AWARE classification: There was poor awareness of the WHO Access, Watch, and Reserve (AWaRe) classification of antibiotics. Only 15% (4/27) had heard about the classification.

Perception of AMR prevalence: All (100%, 27/27) believed AMR to be a problem in Nigeria. However, only 59% (16/27) thought this to be a problem in their health facility; 33% (9/27) were unsure, while 7% (2/27) did not believe AMR to be a problem at their health facility.

Perceptions for tackling AMR: Out of four options for tackling AMR in the society, increasing awareness among patients, enforcing prescription laws, and increased testing for antibiotic susceptibility were jointly the most rated solutions by 96% (26/27), followed by improved training for doctors and pharmacists, 93% (25/27).

Infection Prevention and Control (Hand hygiene): There was a positive attitude to hand hygiene with all (100%, 27/27) affirming the need to perform hand hygiene if they have used gloves in contact with patients or biological material. In contrast, awareness was of the WHO guidance on hand hygiene was moderate. Three-quarters (74%, 20/27) could list the WHO’s five moments of hand hygiene, while the rest were either unsure (22%, 6/27) or could not (4%, 1/27).

Opportunity: The overall score for opportunity was 78%. The majority (85%, 23/27) had easy access to guidelines for managing infections and were confident making antibiotic prescription decisions (81%, 23/27) but did not feel supported to not prescribe antibiotics when not needed, 63% (17/27) (Table 4).

**Table 4.**
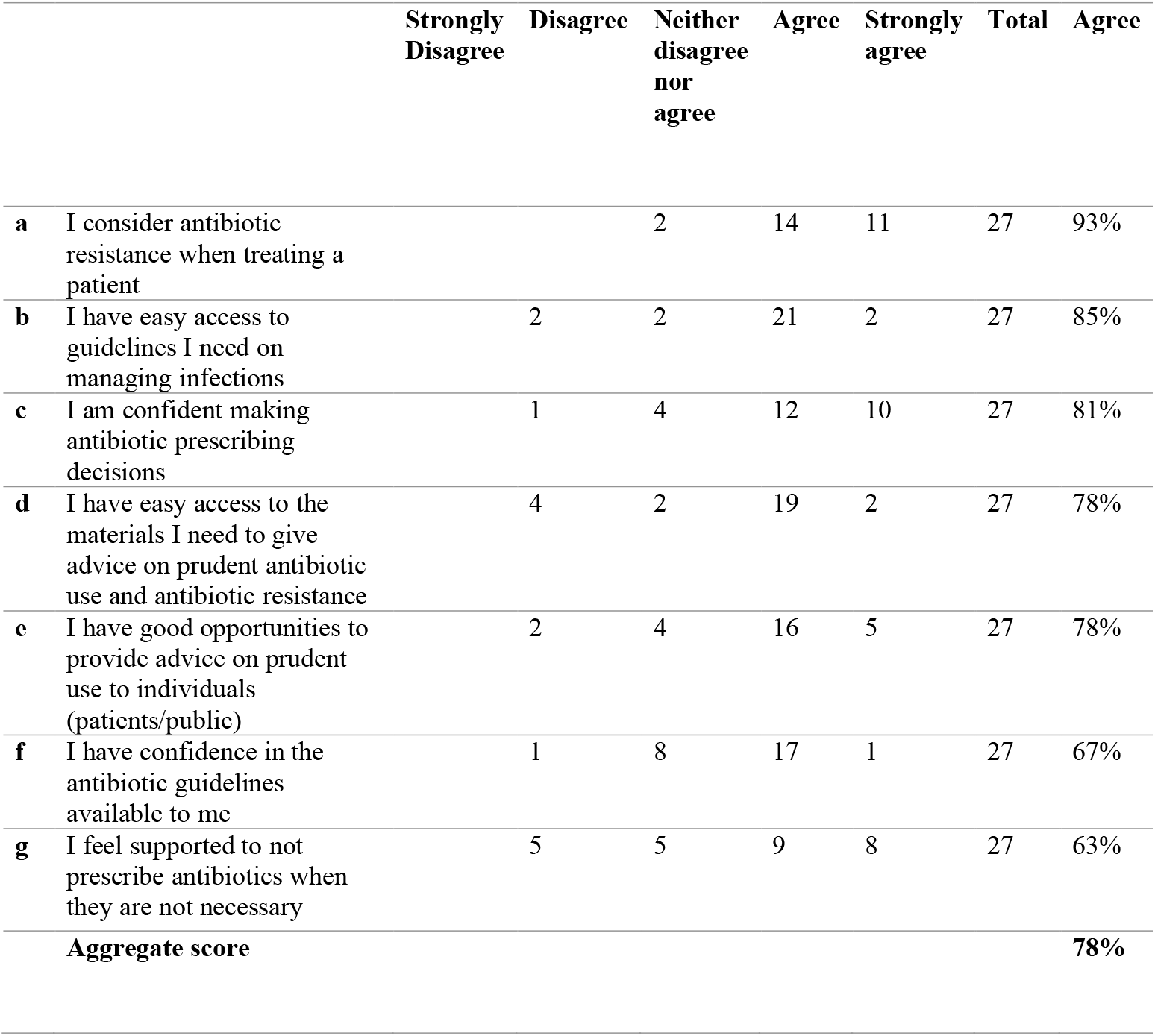
Opportunity for rational (evidence-based) antimicrobial prescribing

The majority reported the presence of a Drug and Therapeutics Committee (74%, 20/27) and a Hospital Formulary or Essential Medicines List (82%, 22/27).

Motivation: The motivation score among participants was 87% overall. Most agreed that there is a connection between their antibiotics prescription pattern and the emergence and spread of AMR in patients (78%, 21/27) and that they have a crucial role in helping to control AMR, 96% (26/27) (Table 5).

**Table 5.** Motivation to improving prescribing by the re-coded variable “agree”

|  |  | **Strongly**<br>**Disagree** | **Disagree** | **Neither**<br>**disagree**<br>**nor**<br>**agree** | **Agree** | **Strongly**<br>**agree** | **Agree** |
| --- | --- | --- | --- | --- | --- | --- | --- |
| **a** | There is a connection between my prescribing of antibiotics and emergence and spread of antibiotic resistant bacteria |  | 1 | 5 | 8 | 13 | 78% |
| **b** | I have a key role in helping control antibiotic resistance |  |  | 1 | 5 | 21 | 96% |
|  | **Aggregate score** |  |  |  |  |  | **87%** |

Note: Aggregate score is the overall average score of participants across questions.

IV. Perceived options to improve antibiotic prescribing: The top measures to improve antibiotic prescribing were: availability of local/national resistance data, availability of local/national guidelines/policies/protocols, readily accessible microbiological data, and easy access to infectious disease physicians (100%, 27/27, each); regular audits and feedback on antibiotic prescribing (96%, 26/27); and educational sessions on prescribing (93%, 25/27). The least rated measures to improve prescribing were computer-aided prescribing (44%, 12/27), restriction of prescription of all antibiotics (30%, 8/27), and speaking to a pharmaceutical representative (15%, 4/27) (Table 6).

**Table 6:**
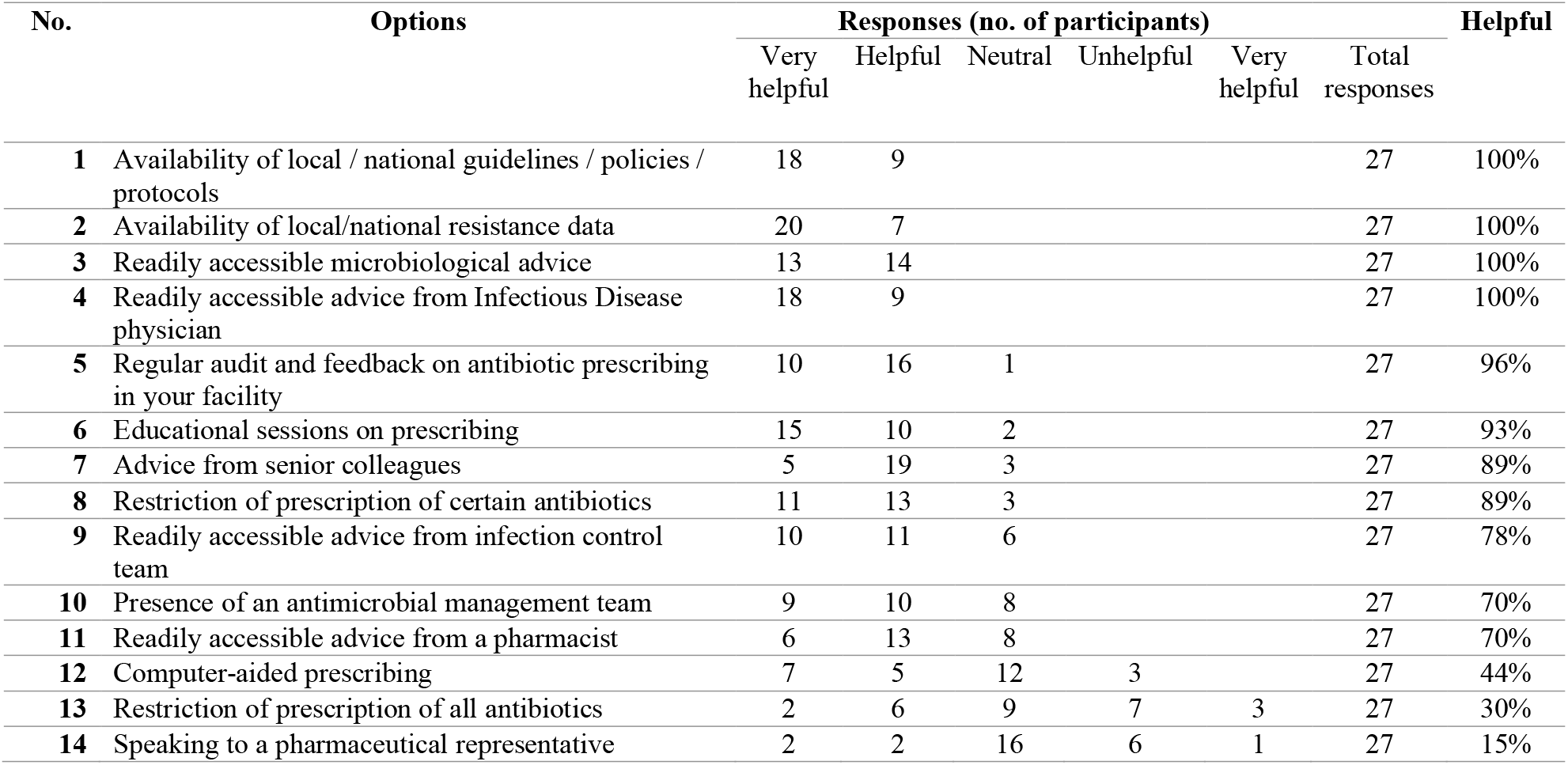
Perceived options among 27 surveyed participants for improving antibiotic prescribing ranked from high to low using the re-coded variable “helpful”

## Discussion

Targeted or evidence-based antibiotic prescribing in children is an urgent clinical need. This is because the indiscriminate or wrong use of antibiotics in children may not only facilitate the emergence and spread of AMR but can also lead to other diseases later in life (29). It is thought that intestinal dysbiosis resulting from early antibiotic exposures predisposes to paediatric idiopathic arthritis, inflammatory bowel disease, asthma, and diabetes(citation).

This study attempted to evaluate the knowledge, attitude, and practices on antimicrobial use, resistance, and stewardship, as well as attitude towards interventions to strengthen AMS among physicians who prescribe antibiotics to children and pregnant women at two tertiary healthcare facilities in Nigeria. While there are studies on antimicrobial utilization in the study location, this only addressed structural gaps and did not focus on children (30).

Empirical prescribing, making antibiotic choices based on experience and not on AST, is the predominant mode of prescribing in this setting, similar to findings previously reported (30). While efforts to change this practice towards more evidence-based prescribing are established or expanding in many countries in the northern hemisphere (31,32), uptake in the southern hemisphere is low. In Nigeria, one study found that none out of 12 surveyed tertiary hospitals had a formal AMS structure (33).

Prescribing is a complex, time-dependent activity in which the prescriber will consider, amongst other things, the symptoms and severity at presentation, the age, allergy history, drug-drug interaction, previous resistant pattern if known, and cost and family dynamics. In almost all cases, the symptoms and severity dictate treatment choice, as is the situation in this study. Without exception, participants in this study recognized the need for resistance data and an antibiotic prescribing guide.

Overall, the antibiotic prescribing pattern in this study agrees with the use pattern across Bayelsa State from a recent prescription audit (30). The most prescribed antibiotics in this study were cefuroxime (a cephalosporin with antistaphylococcal effect), amoxicillin including amoxicillin-clavulanic acid (penicillin), ciprofloxacin (a fluoroquinolone), and azithromycin (a macrolide). These were also the most frequently prescribed in the audit, except for azithromycin. A 20-year literature review of irrational prescribing also found penicillins, fluoroquinolones, and cephalosporins to be the most prescribed antibiotics worldwide (34).

Most of the commonly prescribed antibiotics (azithromycin, cefuroxime, and ciprofloxacin) were in the WHO Watch category - a group of antibiotics of different therapeutic classes that should be prioritized because of their more significant risk of selecting for AMR. One reason for using the AWaRe classification is to ensure that 60% of prescribing is from the access group of antibiotics. The findings from the prescription audit earlier (30) showed that this is not yet the case in this setting. Indeed, most of the prescribers included in this survey had not heard of this categorization intended to improve antibiotic use and reduce AMR (6,35).

Lack of access to rapid pathogen detection and antimicrobial susceptibility testing remains a barrier to AMS. Rapid diagnostic tests (RDTs) can reduce the time interval to use AST for guiding prescribing (36). RDTs using molecular techniques are now commercially available for the identification of various microbial etiological agents, including syphilis (37), invasive Streptococcus pyogenes (38), and helpful in differentiating bacterial from viral causes of respiratory diseases (39). This innovation has also led to several approved devices that can rapidly assess (<1-24hrs) the antimicrobial identification and sensitivity when there is infection (40,41). However, RDTs may not be readily available in resource-constrained settings like ours.

Approaches to tackle AMR in tertiary healthcare facilities should start from the community. The lack of suitable formulations for children potentially complicates antimicrobial use. One study of the WHO Essential Medicines List used to inform the selection, and use of medicines for children found that over 70% of the recommended medicines for oral use in the 2011-2019 lists were not suitable for use in children less than 5 years old (42). This situation leads to the extemporaneous preparation or compounding of medicines for children with potential safety and efficacy issues. Although inpatient antibiotic use is usually via the parenteral route for younger children (43,44), the easy over-the-counter (OTC) access to oral antibiotics at the community level can complicate efforts aimed at AMS. It is, therefore, important to address AMR through community-targeted programs in order to reduce the burden placed on tertiary healthcare facilities. A recent study suggests that reconstituted amoxicillin-clavulanic acid suspension stored under room temperature these settings undergoes rapid quality changes over 7 days (45), making addressing paediatric AMS challenging.

This study clearly shows that there is a need to improve providers’ knowledge on AMR and AMS. Providers also need to be better acquainted with the WHO guidelines among pediatricians and other practitioners who prescribe antibiotics for use in the pediatric population. Continuing medical education for the rational use of antibiotics at regular intervals could motivate the necessary behaviour change among pediatric practitioners. This study is not without its limitations. For one, we did not assess the impact of cost on prescribing behavior, nor did it evaluate practitioners’ knowledge of the pharmacodynamics and pharmacokinetics of antibiotics prescribed, as this was beyond the scope of the current research effort.

## Conclusion

This study evaluated the knowledge, attitude, and practices of providers who prescribe antibiotics to children at two tertiary healthcare facilities in Bayelsa State, Nigeria, with the goal of understanding barriers and facilitators for establishing an antimicrobial stewardship program for children.

There is a need for readily accessible resistance data and information to guide rational antimicrobial prescribing in tertiary healthcare settings in the Niger Delta region of Nigeria. A clinical decision support system that integrates local resistance data to guide antibiotic prescribing would be an effective pediatric AMS strategy in this and similar settings. To be effective, this system should be readily accessible to prescribers and regularly updated with resistance data. Shared ownership, fostered through a participatory approach, could help enhance uptake and use. Rapid diagnostic tests to rapidly inform antibiotic choice are a must and would facilitate transitioning from empiric therapy to definitive therapy. Future research should focus on improving AMS at the institutional and national levels through the systematic collection and use of AMR data.

## Data Availability

All data produced in the present work are contained in the manuscript

## Disclosures regarding real or perceived conflicts of interest

None to declare.

