## Supplemental Table 1 for "Antimicrobial stewardship among Nigerian children: A pilot study of the knowledge, attitude, and practices of prescribers at two tertiary healthcare facilities in Bayelsa State"

**Supplement Table 1. Questionnaire_Survey_prescribers_NDUTH_Nigeria_Jul26_2021**

1. **Have you prescribed an antibiotic within the past 3 months?**

   ○ Yes

   ○ No

2. **How many patients do you see on a daily basis?**

   ○ Less than 25

   ○ Between 25-49

   ○ Between 50-100

   ○ Over 100

3. **Among the patients you see, approximately what percentage are children under 12 years old?**

   ○ less than 10%

   ○ between 10-24%

   ○ between 25-50%

   ○ over 50%

4. **How many prescriptions for antibiotics have you written in the last 7 days?**

   ○ Less than 50

   ○ Between 50-99

   ○ Between 100-149

   ○ Over 150

5. **To what extent do you agree or disagree with each of the following statements:**

| | Strongly Disagree | Disagree | Neither disagree nor agree | Agree | Strongly agree |
|---|---|---|---|---|---|
| • I know what antibiotic resistance is | ☐1 | ☐2 | ☐3 | ☐4 | ☐5 |
| • I know what information to give to individuals about prudent use of antibiotics and antibiotic resistance | ☐1 | ☐2 | ☐3 | ☐4 | ☐5 |
| • I know sufficient knowledge about how to use antibiotics appropriately for my current practice | ☐1 | ☐2 | ☐3 | ☐4 | ☐5 |

6. **Please answer whether you believe each of these statements to be True (T) or False (F):**

| | **True** | **False** |
|---|---|---|
| • **Antibiotics are effective against viruses** | ☐ | ☐ |
| • **Antibiotics are effective against cold and flu** | ☐ | ☐ |
| • **Unnecessary use of antibiotics makes them become ineffective** | ☐ | ☐ |
| • **Taking antibiotics has associated side effects such as diarrhea, colitis, or allergy** | ☐ | ☐ |
| • **Every person treated with antibiotics is at an increased risk of antibiotic resistant infection** | ☐ | ☐ |
| • **Antibiotic resistant bacteria can spread from person to person** | ☐ | ☐ |
| • **Healthy people can carry antibiotic resistant bacteria** | ☐ | ☐ |

7. **Do you think antibiotic resistance is a problem in Nigeria?**

○ Yes

○ No

○ Unsure

**8. Do you think antibiotic resistance is a problem in your health care facility?**

○ Yes

○ No

○ Unsure

**9. How do you suggest the problem of antibiotic resistance can be tackled in Nigeria? (Choose ALL that apply)**

○ Increase testing for antibiotic susceptibility

○ Increase awareness among patients

○ Enforce prescription laws for the dispensing of antibiotics

○ Improved training of doctors

○ Other____________________________________________________________

**10. To what extent do you agree or disagree that the following environmental and animal health factors are important in contributing to antibiotic resistance in bacteria from humans?**

| | Strongly Disagree | Disagree | Neither disagree nor agree | Agree | Strongly agree |
|---|---|---|---|---|---|
| • Environmental factors such as waste water in the environment | ☐1 | ☐2 | ☐3 | ☐4 | ☐5 |
| • Excessive use of antibiotics in livestock and food production | ☐1 | ☐2 | ☐3 | ☐4 | ☐5 |

**11. To what extent do you agree or disagree with each of the following statements:**

| | Strongly Disagree | Disagree | Neither disagree nor agree | Agree | Strongly agree |
|---|---|---|---|---|---|
| • I have easy access to guidelines I need on managing infections | ☐1 | ☐2 | ☐3 | ☐4 | ☐5 |
| • I have easy access to the materials I need to give advice on prudent antibiotic use and antibiotic resistance | ☐1 | ☐2 | ☐3 | ☐4 | ☐5 |
| • I have good opportunities to provide advice on prudent use to individuals (patients/public) | ☐1 | ☐2 | ☐3 | ☐4 | ☐5 |
| • I am confident making antibiotic prescribing decisions | ☐1 | ☐2 | ☐3 | ☐4 | ☐5 |
| • I have confidence in the antibiotic guidelines available to me | ☐1 | ☐2 | ☐3 | ☐4 | ☐5 |
| • I consider antibiotic resistance when treating a patient | ☐1 | ☐2 | ☐3 | ☐4 | ☐5 |
| • I feel supported to not prescribe antibiotics when they are not necessary | ☐1 | ☐2 | ☐3 | ☐4 | ☐5 |

**12. To what extent do you agree or disagree with the following statements:**

| | Strongly Disagree | Disagree | Neither disagree nor agree | Agree | Strongly agree |
|---|---|---|---|---|---|
| • There is a connection between my prescribing of antibiotics and emergence and spread of antibiotic resistant bacteria | ☐1 | ☐2 | ☐3 | ☐4 | ☐5 |
| • I have a key role in helping control antibiotic resistance | ☐1 | ☐2 | ☐3 | ☐4 | ☐5 |

**13. Can you list the WHO's five moments of hand hygiene?**

○ Yes

○ No

○ Unsure

**14. Do you need to perform hand hygiene (as often as recommended) if you have used gloves in contact with patients or biological material?**

○ Yes

○ No

○ Unsure

**15. In your facility, do you follow Standard Treatment Guidelines (or a guide specific to prescribing antibiotics)?**

○ Yes

○ No

**16. Does your facility have a Drug and Therapeutic Committee?**

○ Yes

○ No

**17. a. Is there a Hospital Formulary or an Essential Medicine List?**

- ◯ Yes
- ◯ No

**b. If yes, which**:

- ◯ Hospital formulary
- ◯ Hospital or National Essential Medicine List
- ◯ Both

**18. a. Is it possible to perform laboratory antibiotic susceptibility testing on-site or off-site?**

- ◯ Yes
- ◯ No
- ◯ Not applicable

**b. If yes, is it on-site or off-site?**

- ◯ On-site
- ◯ Off-site

**19. How many patients, on average, do you see in a month?**

- ◯ 0 – 99
- ◯ 100 – 199
- ◯ 200 – 299
- ◯ 300 or more

**20. How often do you see treatment failure within a month (no response to therapy, switch to new antibiotic, or addition of new antibiotic)?**

- ◯ Less than 10%
- ◯ Between 11 – 25%
- ◯ Between 26 – 50%
- ◯ 50% or more

**21. a. Are antibiotic susceptibility tests performed before antibiotic selection?**

○ Yes

○ No

**b. If not, why? (Please, check all that apply)**

○ Availability

○ Time pressure

○ Other. Please specify________________________________________________

**c. If yes, how often?**

○ Always

○ Frequently

○ Infrequently

**d. If yes, in what cases are antibiotic susceptibility tests usually used? (List up to three)**

______________________________________________________________________________

______________________________________________________________________________

______________________________________________________________________________

**22. Please select the top three antibiotics that you usually prescribe?**

| | First Choice | Second Choice | Third Choice |
|---|---|---|---|
| • Amoxicillin (including amoxicillin-clavulanate) | ☐ | ☐ | ☐ |
| • Azithromycin | ☐ | ☐ | ☐ |
| • Cefuroxime | ☐ | ☐ | ☐ |
| • Ciprofloxacin | ☐ | ☐ | ☐ |
| • Co-trimoxazole | ☐ | ☐ | ☐ |
| • Metronidazole | ☐ | ☐ | ☐ |
| • Other: (Please list) ______________________ | ☐ | ☐ | ☐ |
| • Other: (Please list) ______________________ | ☐ | ☐ | ☐ |
| • Other: (Please list) ______________________ | ☐ | ☐ | ☐ |

**23. What is your main consideration for prescribing antibiotics? (Please, specify all that apply)**

- ○ Antibiotic susceptibility testing information
- ○ Symptoms of patient
- ○ Clinical severity of case
- ○ Other: Please, specify ______________________________________

**24. What are the top three things you considered for antibiotic CHOICE when you prescribe?**

| | First | Second | Third |
|---|---|---|---|
| • **Availability of antibiotic in the facility** | ☐ | ☐ | ☐ |
| • **Antibiotic susceptibility testing information** | ☐ | ☐ | ☐ |
| • **Broad-spectrum nature of antibiotic** | ☐ | ☐ | ☐ |
| • **Symptoms of patient** | ☐ | ☐ | ☐ |
| • **Other: (Please list) ______________________** | ☐ | ☐ | ☐ |
| • **Other: (Please list) ______________________** | ☐ | ☐ | ☐ |
| • **Other: (Please list) ______________________** | ☐ | ☐ | ☐ |

**25. When you prescribe antibiotics for a patient, do you usually prescribe? [Combination products count as 1]**

◯ 1

◯ 2-3

◯ >3

**26. Please indicate which GROUPs are MOST FREQUENTLY prescribed for the following infections?**

| | RTI | UTI | GIT Diseases | Dental Infections | Skin and Soft Tissue infections |
|---|---|---|---|---|---|
| • Penicillins | ☐1 | ☐2 | ☐3 | ☐4 | ☐5 |
| • Cephalosporins | ☐1 | ☐2 | ☐3 | ☐4 | ☐5 |
| • Fluoroquinolones | ☐1 | ☐2 | ☐3 | ☐4 | ☐5 |
| • Macrolides | ☐1 | ☐2 | ☐3 | ☐4 | ☐5 |
| • Trimethoprim | ☐1 | ☐2 | ☐3 | ☐4 | ☐5 |
| • Others (please specify): ___________ | ☐1 | ☐2 | ☐3 | ☐4 | ☐5 |

**27. Do you feel pressured to prescribe antibiotics?**

○ Yes. If yes, why? ______________________________________________________________

○ No

**28. Are any of these reasons that you feel pressured to prescribed antibiotics:**

| | Strongly Disagree | Disagree | Neither disagree nor agree | Agree | Strongly agree |
|---|---|---|---|---|---|
| • Patient demand | ☐1 | ☐2 | ☐3 | ☐4 | ☐5 |
| • Condition of patient | ☐1 | ☐2 | ☐3 | ☐4 | ☐5 |
| • Fear of complications | ☐1 | ☐2 | ☐3 | ☐4 | ☐5 |
| • Need to provide rapid relief | ☐1 | ☐2 | ☐3 | ☐4 | ☐5 |

Please specify any other reasons that you feel pressured to prescribe antibiotics:

___________________________________

**29. Do you usually prescribe antibiotics by generic or brand name?**

○ Generic

○ Brand

**30. If you prescribe by brand, what are your reasons for doing so? (Please, specify all that apply)**

○ Clinical efficacy (high bacterial resistance to generics)

○ Better quality

○ Other (please specify) ___________________________________________________________________

**31. To optimize the use of antibiotics, the WHO has classified antibiotics as Access, Watch, & Reserve.**

**a. Have you heard about these categories?**

◯ Yes

◯ No

**b. If yes, do they affect your prescribing?**

◯ Yes

◯ No

**32. What measures do you think would be the most helpful in improving antibiotic prescribing?**

| | Very helpful | Helpful | Neutral | Unhelpful | Very unhelpful |
|---|---|---|---|---|---|
| • Educational sessions on prescribing | ☐1 | ☐2 | ☐3 | ☐4 | ☐5 |
| • Availability of local / national guidelines / policies / protocols | ☐1 | ☐2 | ☐3 | ☐4 | ☐5 |
| • Availability of local/national resistance data | ☐1 | ☐2 | ☐3 | ☐4 | ☐5 |
| • Computer-aided prescribing | ☐1 | ☐2 | ☐3 | ☐4 | ☐5 |
| • Presence of an antimicrobial management team | ☐1 | ☐2 | ☐3 | ☐4 | ☐5 |
| • Readily accessible microbiological advice | ☐1 | ☐2 | ☐3 | ☐4 | ☐5 |
| • Readily accessible advice from Infectious Disease physician | ☐1 | ☐2 | ☐3 | ☐4 | ☐5 |
| • Readily accessible advice from a pharmacist | ☐1 | ☐2 | ☐3 | ☐4 | ☐5 |

| | | | | | |
|---|---|---|---|---|---|
| • Readily accessible advice from infection control team | ☐1 | ☐2 | ☐3 | ☐4 | ☐5 |
| • Advice from senior colleagues | ☐1 | ☐2 | ☐3 | ☐4 | ☐5 |
| • Speaking to a pharmaceutical representative | ☐1 | ☐2 | ☐3 | ☐4 | ☐5 |
| • Restriction of prescription of certain antibiotics | ☐1 | ☐2 | ☐3 | ☐4 | ☐5 |
| • Restriction of prescription of **all** antibiotics | ☐1 | ☐2 | ☐3 | ☐4 | ☐5 |
| • Regular audit and feedback on antibiotic prescribing in your PHC | ☐1 | ☐2 | ☐3 | ☐4 | ☐5 |

**33. How many years of practice experience do you have?**

○ Less than 1 year

○ 1 – 5 years

○ 6 – 10 years

○ 11 – 15 years

○ 15 years or more

**34. What is your specialty?**

○ Medicine

○ Surgery

○ Pediatrics

○ Anesthetics

○ Obstetrics / Gynecology

○ Psychiatry

○ Other

**35. Do you have additional comments you find relevant?**

______________________________________________________________________________

______________________________________________________________________________

______________________________________________________________________________

**Thank you very much for taking part in this survey!**
