## Supplemental Table 2 for "Antimicrobial stewardship among Nigerian children: A pilot study of the knowledge, attitude, and practices of prescribers at two tertiary healthcare facilities in Bayelsa State"

**Supplement Table 2 Mapping of questionnaire onto COM-B components**

| Section | Questions | Response |
|---|---|---|
| **I. Demographic characteristics** | 1) How many years of practice experience do you have?<br>2) What is your specialty? | Multiple choice |
| **II. Prescribing behavior/practices** | 1) Have you prescribed an antibiotic within the past 3 months?<br>2) How many patients do you see on a daily basis?<br>3) Among the patients you see, approximately what %age are children <12 years old?<br>4) How many prescriptions for antibiotics have you written in the last 7 days?<br>5) How many patients, on average, do you see in a month?<br>6) a. Are antibiotic susceptibility tests performed before antibiotic selection?<br>b. If not, why? (Please, check all that apply) [Availability, Time pressure, Other ____<br>c.If yes, how often?<br>d.If yes, in what cases are antibiotic susceptibility tests usually used? (List up to three)<br>7) What is your main consideration for prescribing antibiotics? (Specify all that apply)<br>○ Antibiotic susceptibility tests/Microbial Culture and Sensitivity (MCS) information<br>○ Symptom of patient<br>○ Clinical severity of case<br>○ Patient demand<br>8) a. Do you feel pressured to prescribe antibiotics?<br>b. If yes, are any of these reasons that you feel pressured to prescribe antibiotics:<br>○ Patient demand<br>○ Condition of patient<br>○ Fear of complications<br>○ Need to provide rapid relief<br>○ Others __________________<br>9) Do you usually prescribe antibiotics by generic or brand name? | Yes/No<br><br>Yes/No<br><br>5-point Likert Scale (from left to right; Strongly disagree to Strongly agree) |

<table>
<tr>
<td></td>
<td>10) If you prescribe by brand, what are your reasons for doing so? (Specify all that apply)<br><br>o Clinical efficacy (high bacterial resistance to generics)<br>o Better quality<br>o Other ____________<br><br>11) What are the top three things you considered for antibiotic CHOICE when you prescribe?<br><br>o Availability of antibiotic in the facility<br>o Antibiotic susceptibility testing information<br>o Broad-spectrum nature of antibiotic<br>o Symptoms of patient<br>o Others: (Please list) ___________________<br><br>12) What is your main consideration for prescribing antibiotics? (Specify all that apply)<br><br>o Antibiotic susceptibility testing information<br>o Symptoms of patient<br>o Clinical severity of case<br>o Other: Please, specify _________________</td>
<td></td>
</tr>
<tr>
<td>Antibiotic prescribing pattern</td>
<td>1) Please select the top three antibiotics that you usually prescribe?<br><br>o Amoxicillin (including amoxicillin-clavulanate)<br>o Azithromycin<br>o Cefuroxime<br>o Ciprofloxacin<br>o Co-trimoxazole<br>o Metronidazole<br>o Others: (Please list) _____________________<br>2) When you prescribe antibiotics for a patient, do you usually prescribe? [Combination products count as 1] [1, 2-3, >3]<br><br>3) Please indicate which GROUPs are MOST FREQUENTLY prescribed for the following infections?<br><br>
<table>
<tr><td></td><td>RTI</td><td>UTI</td><td>GIT Diseases</td><td>Dental Infections</td><td>Skin & Soft Tissue infections</td></tr>
<tr><td>• Penicillins</td><td></td><td></td><td></td><td></td><td></td></tr>
<tr><td>• Cephalosporins</td><td></td><td></td><td></td><td></td><td></td></tr>
<tr><td>• Fluoroquinolones</td><td></td><td></td><td></td><td></td><td></td></tr>
<tr><td>• Macrolides</td><td></td><td></td><td></td><td></td><td></td></tr>
</table>
</td>
<td></td>
</tr>
</table>

| | | |
|---|---|---|
| | • Trimethoprim<br>• Other (please specify) | |
| **II Capacity, Opportunity, and Motivation** | *Capacity – Knowledge*<br>1) To what extent do you agree or disagree with each of the following statements:<br>o I know what antibiotic resistance is<br>o I know what information to give to individuals about prudent use of antibiotics and antibiotic resistance<br>o I know sufficient knowledge about how to use antibiotics appropriately for my current practice<br><br>2) Please answer whether you believe each of these statements to be True or False:<br>o Antibiotics are effective against viruses<br>o Antibiotics are effective against cold and flu<br>o Unnecessary use of antibiotics makes them become ineffective<br>o Taking antibiotics has associated side effects such as diarrhea, colitis, or allergy<br>o Every person treated with antibiotics is at an increased risk of antibiotic-resistant infection<br>o Antibiotic resistant bacteria can spread from person to person<br>o Healthy people can carry antibiotic resistant bacteria<br><br>3) To what extent do you agree or disagree that the following environmental and animal health factors are important in contributing to antibiotic resistance in bacteria from humans?<br>o Environmental factors such as waste water in the environment<br>o Excessive use of antibiotics in livestock and food production<br><br>4) Do you need to perform hand hygiene (as often as recommended) if you have used gloves in contact with patients or biological material?<br>5) Can you list the WHO's five moments of hand hygiene? | 5-point Likert Scale (from left to right; Strongly disagree to Strongly agree)<br><br>True or False<br><br>5-point Likert Scale (from left to right; Strongly disagree to Strongly agree)<br><br>Yes/No |
| | *Capacity - Awareness & Perceptions*<br>1) Do you think antibiotic resistance is a problem in Nigeria? | Yes/No |

| | | |
|---|---|---|
| | 2) Do you think antibiotic resistance is a problem in your healthcare facility?<br><br>3) How do you suggest the problem of antibiotic resistance can be tackled in Nigeria? (Choose ALL that apply)<br>o Increase testing for antibiotic susceptibility<br>o Increase awareness among patients<br>o Enforce prescription laws for the dispensing of antibiotics<br>o Improved training of doctors and pharmacists<br>o Other___________________________________________<br><br>4) To optimize the use of antibiotics, the WHO has classified antibiotics as Access, Watch, & Reserve. Have you heard about these categories?<br><br>5) If yes, do they affect your prescribing?<br><br>6) How often do you see treatment failure within a month (no response to therapy, switch to new antibiotic, or addition of new antibiotic)? | Yes/No<br><br><br><br><br><br><br><br>Yes/No<br><br>Yes/No |
| | *Motivation*<br><br>1. To what extent do you agree or disagree with the following statements:<br>○ There is a connection between my prescribing of antibiotics and emergence and spread of antibiotic resistant bacteria<br>○ I have a key role in helping control antibiotic resistance | 5-point Likert Scale (from left to right; Strongly disagree to Strongly agree) |
| | *Opportunity – Attitude or confidence in prescribing and awareness of AMS structures*<br><br>1) To what extent do you agree or disagree with each of the following statements:<br><br>○ I have easy access to guidelines I need on managing infections<br>○ I have easy access to the materials I need to give advice on prudent antibiotic use and antibiotic resistance<br>○ I have good opportunities to provide advice on prudent use to individuals (patients/public)<br>○ I am confident making antibiotic prescribing decisions<br>○ I have confidence in the antibiotic guidelines available to me<br>○ I consider antibiotic resistance when treating a patient<br>○ I feel supported to not prescribe antibiotics when they are not necessary | 5-point Likert Scale (from left to right; Strongly disagree to Strongly agree) |

| | | |
|---|---|---|
| | 2) In your facility, do you follow Standard Treatment Guidelines (or a guide specific to prescribing antibiotics)?<br>3) Does your facility have a Drug and Therapeutic Committee (DTC)?<br>4) If yes, which? [Hospital Formulary; Hospital of National Essential Medicines List; Both]<br>5) Is it possible to perform laboratory antibiotic susceptibility testing on-site or off-site?<br>6) If yes, is it on-site or off-site? | Yes / No<br><br>Yes / No |
| **III. Perceived strategies to improve prescribing** | 1) What measures do you think would be the most helpful in improving antibiotic prescribing?<br>o Educational sessions on prescribing<br>o Availability of local / national guidelines / policies / protocols<br>o Availability of local/national resistance data<br>o Computer-aided prescribing<br>o Presence of an antimicrobial management team<br>o Readily accessible microbiological advice<br>o Readily accessible advice from Infectious Disease physician<br>o Readily accessible advice from a pharmacist<br>o Readily accessible advice from infection control team<br>o Advice from senior colleagues<br>o Speaking to a pharmaceutical representative<br>o Restriction of prescription of certain antibiotics<br>o Restriction of prescription of all antibiotics<br>o Regular audit and feedback on antibiotic prescribing in your PHC | 5-point Likert Scale (from left to right; Very helpful to Very unhelpful) |

Notes: Items with lettered options (a, b, c) representing adaptive questions were counted as individual items in the text. Hence the total number of questions in this table is less than reported in the manuscript.
