## Supplemental Table 3 for "Antimicrobial stewardship among Nigerian children: A pilot study of the knowledge, attitude, and practices of prescribers at two tertiary healthcare facilities in Bayelsa State"

Supplement Table 3a Summary of results for responses with frequency counts. [Other results presented in text, and Tables 3b&c below]

| | | n | % (n/27) |
|---|---|---|---|
| **1** | Consent | | |
| | Yes | 27 | 100% |
| | No | 0 | |
| **2** | Prescribed antibiotic past 3 months | | |
| | Yes | 27 | 100% |
| | No | 0 | |
| **3** | Patients, daily load | | |
| | <25 | 21 | 78% |
| | 25-50 | 4 | 15% |
| | 50-100 | 1 | 4% |
| | >100 | 1 | 4% |
| **4** | Clientele under 12 | | |
| | <10% | 3 | 11% |
| | 10-<25% | | |
| | 25-50% | 6 | 22% |
| | >50% | 18 | 67% |
| **5** | Antibiotic prescriptions last 7 days | | |
| | <50 | 23 | 85% |
| | 50-99 | 4 | 15% |
| | 100-149 | | |
| | >150 | | |
| **6** | Antimicrobial Resistance (AMR) a problem in Nigeria | | |
| | Yes | 27 | 100% |
| | No | | |
| **7** | AMR a problem at your facility | | |
| | Yes | 16 | 59% |
| | No | 2 | 7% |
| | Unsure | 9 | 33% |
| **8** | Tackling AMR in society | | |
| | Increased testing | 26 | 96% |
| | Increase patient awareness | 26 | 96% |
| | Enforce prescription laws | 26 | 96% |
| | Improved training for doctors and pharmacists | 25 | 93% |
| **9** | WHO's five moments of hand hygiene | | |
| | Yes | 20 | 74% |
| | No | 1 | 4% |
| | Unsure | 6 | 22% |
| **10** | Hand hygiene after using gloves | | |

| | | | |
|---|---|---|---|
| | Yes | 27 | 100% |
| | No | | |
| | Unsure | | |
| **11** | Use of Standard Treatment Guidelines (STG) at facility | | |
| | Yes | 15 | 56% |
| | No | 10 | 37% |
| | Unsure | 2 | 7% |
| **12** | Drug and Therapeutic Committee (DTC) at facility | | |
| | Yes | 20 | 74% |
| | No | 6 | 22% |
| **13** | a. Hospital Formulary or Essential Medicines List (EML) | | |
| | Yes | 24 | 89% |
| | No | 3 | 11% |
| | b. If Yes, which | | |
| | Formulary | 9 | 33% |
| | EML | 8 | 30% |
| | Both | 5 | 19% |
| **14** | a. Possible to perform Antibiotic Susceptibility Testing (AST) onsite/offsite | | |
| | Yes | 18 | 67% |
| | No | 2 | 7% |
| | Not applicable | 7 | 26% |
| | b. If yes, is it on-site or off-site | | |
| | On-site | 16 | 59% |
| | Off-site | 2 | 7% |
| **15** | Patients, monthly load | | |
| | 0-99 | 19 | 70% |
| | 100-199 | 5 | 19% |
| | 200-299 | 2 | 7% |
| | >300 | 1 | 4% |
| **16** | Treatment failure within a month | | |
| | <10% | 14 | 52% |
| | 11-25% | 9 | 33% |
| | 26-50% | 3 | 11% |
| | >50% | 1 | 4% |
| **17** | a. AST before antibiotic selection | | |
| | Yes | 3 | 11% |
| | No | 24 | 89% |
| | b. If not, why? | | |
| | Availability | 12 | 44% |
| | Time pressure | 19 | 70% |
| | Other | | |
| | Finance | 2 | 7% |
| | Very ill patient | 4 | 15% |
| | c. If yes, how often are AST performed | | |

| | | | |
|---|---|---|---|
| | Always | | |
| | Frequently | 2 | 7% |
| | Infrequently | 6 | 22% |
| | d. If yes, cases when AST are used | | |
| | UTI in pregnancy, wound sepsis, puerperal sepsis, post-abortion sepsis, persistent fever unresponsive to treatment, bronchopneumonia, meningitis, otitis media | | |
| **18** | Top three antibiotics prescribed | | |
| | Amoxicillin (including amoxicillin-clavulanate) | | |
| | First choice | 11 | 41% |
| | Second choice | 8 | 30% |
| | Third choice | 3 | 11% |
| | Azithromycin | | |
| | First choice | 1 | 4% |
| | Second choice | 6 | 22% |
| | Third choice | 9 | 33% |
| | Cefuroxime | | |
| | First choice | 11 | 41% |
| | Second choice | 5 | 19% |
| | Third choice | 8 | 30% |
| | Ciprofloxacin | | |
| | First choice | 1 | 4% |
| | Second choice | 6 | 22% |
| | Third choice | 4 | 15% |
| | Cotrimoxazole | | |
| | First choice | | 0% |
| | Second choice | | 0% |
| | Third choice | | 0% |
| | Metronidazole | | 0% |
| | First choice | | 0% |
| | Second choice | | 0% |
| | Third choice | | 0% |
| **19** | Main consideration for prescribing antibiotics | | |
| | Antibiotic susceptibility testing information | 16 | 59% |
| | Symptoms of patient | 22 | 81% |
| | Clinical severity of case | 22 | 81% |
| | Other | | |
| | Assumed prior antibiotic use before referral | 2 | 7% |
| **20** | Top three considerations for antibiotic choice | | |
| | Availability of antibiotic at facility | | |
| | First choice | 4 | |
| | Second choice | 1 | |
| | Third choice | 7 | |
| | Antibiotic susceptibility testing information | | |

| | | | |
|---|---|---|---|
| | First choice | 6 | |
| | Second choice | 2 | |
| | Third choice | 1 | |
| | Broad-spectrum nature of antibiotic | | |
| | First choice | 1 | |
| | Second choice | 11 | |
| | Third choice | 8 | |
| | Symptoms of patient | | |
| | First choice | 13 | |
| | Second choice | 4 | |
| | Third choice | 3 | |
| | Standard Treatment Guidelines/Formulary/EML | | |
| | First choice | 3 | |
| | Second choice | 7 | |
| | Third choice | 1 | |
| | Others - Not listed; or not already captured | | |
| | Safety range | 1 | |
| | Cost/affordability | 3 | |
| **21** | No of antibiotics usually prescribed | | |
| | One | | |
| | Two-three | 10 | 37% |
| | More than 3 | | |
| **22** | Pressure to prescribe antibiotics | | |
| | Yes | 15 | 56% |
| | No | 12 | 44% |
| **23** | a. Prescribing | | |
| | Brand | 8 | 30% |
| | Generic | 19 | 70% |
| | b. Reasons for brand prescribing (select all that apply) | | |
| | Clinical efficacy (high bacterial resistance to generics) | 5 | 19% |
| | Better quality | 9 | 33% |
| | Others [severity of the symptoms] | | |
| **24** | a. Heard about WHO AWARE categories | | |
| | Yes | 4 | 15% |
| | No | 23 | 85% |
| | b. If yes, do they affect your prescribing behavior | | |
| | Yes | 2 | 7% |
| | No | 25 | 93% |
| **25** | Years of practice experience | | |
| | 1-5 years | 4 | 15% |
| | 6-10 years | 10 | 37% |
| | 11-15 years | 7 | 26% |
| | >15 years | 6 | 22% |

*Table 3b Antibiotic classes prescribed by survey participants (n=27) for specific disease groups*

| | Penicillins | Cephalo-sporins | Fluoro-quinolones | Macrolides | Sulfonamide/ Trimethoprim |
|---|---|---|---|---|---|
| **Respiratory Tract Infections** | 81% | 33% | 7% | 78% | 56% |
| **Urinary Tract Infections** | 30% | 33% | 63% | 11% | 4% |
| **Gastro-intestinal Diseases** | | 22% | 67% | 30% | 4% |
| **Dental Infections** | 37% | 11% | | 4% | |
| **Skin & Soft Tissue infections** | 44% | 56% | | 4% | 4% |

*Table 3c Reasons for pressure among participants (n=15) who felt pressure to prescribe antibiotics*

| | Strongly disagree | Disagree | Neither disagree nor agree | Agree | Strongly agree | "Agree" |
|---|---|---|---|---|---|---|
| Patient demand | 3 | 4 | 2 | 2 | 4 | 40% |
| Condition of patient | | | | 10 | 5 | 100% |
| Fear of complications | | 1 | | 8 | 6 | 93% |
| Need to provide rapid relief | | 3 | 3 | 5 | 3 | 53% |
